# Systemic immune mediators predict therapeutic response and tumor-infiltrating lymphocyte intensity in triple-negative breast cancer

**DOI:** 10.1101/2020.10.15.20212597

**Authors:** Ananda D. Lopes, Nayane A.L. Galdino, Amanda B. Figueiredo, Rafael C. Brianese, Katia L.P. Morais, Marina De Brot, Cynthia A.B.T, Osório, Andrea Teixeira-Carvalho, Vinicius F. Calsavara, Guilherme F.B. Evangelista, Natalia S. Alves, Fabiana B. Makdissi, Solange M. Sanches, Vladmir C. Cordeiro de Lima, Dirce M. Carraro, Kenneth J. Gollob

## Abstract

Triple-negative breast cancer (TNBC) is an aggressive form of breast cancer defined by the lack of expression of estrogen receptor, progesterone receptors, and of the human epithelial growth factor receptor 2. Neoadjuvant chemotherapy has proven efficacy in the treatment of TNBC, and a pathological complete response (pCR) is predictive of improved long-term survival. The immune response exerts a vital role in response to neoadjuvant chemotherapy, as indicated by the relation between the percentage of stromal tumor-infiltrating lymphocytes (TILs) in pre-treated tumor tissue samples and the likelihood of achieving pCR. Despite this, the relationship between the systemic immune response and the tumor microenvironment is unclear. In this prospective study, we determined the systemic plasma immune profile of TNBC patients before treatment using a panel of 27 immune mediators and measured the percentage of TILs from the same patients. Patients who demonstrated pCR had significantly higher systemic immune mediators; GM-CSF, FGF-basic, VEGF, IL-2, and IL-5, than the non-responders. Moreover, responders displayed a strong positive correlation between the cytokines IFN-γ and IL-7 with the percentage of TILs, while non-responders had a negative or no correlation. Finally, systemic immune mediator levels before treatment predict pCR (AUC range 0.64 - 0.71), and the combination of immune mediators and TILs improved pCR prediction (AUC 0.71 - 0.82). In conclusion, increased systemic immune mediators reflect increased TILs percentage and act as potential predictive biomarkers of pCR for TNBC patients submitted to neoadjuvant chemotherapy.

## Introduction

Breast cancer (BC) is a heterogeneous disease with distinct clinical behavior (1). It is the most prevalent cancer among women globally (excluding non-melanoma skin cancer), affecting approximately 60,000 women in Brazil annually (2). Triple-negative breast cancer (TNBC) is characterized by the lack of expression of estrogen receptor (ER), progesterone receptor (PR), and the human epithelial growth factor receptor 2 (identified as HER2 score 0 or 1+ by immunohistochemistry and no gene amplification by in situ hybridization of HER2 with a score 2+ for ambiguous cases) (3–5), and accounts for 10% to 17% of all invasive carcinomas of the breast (6). TNBC is often associated with more aggressive behavior, a higher frequency of distant metastasis, and a dismal prognosis (7).

Taxanes and anthracycline are the most used neoadjuvant therapy regimens for breast cancer, and approximately 30-40% of patients treated with these drugs obtain pathological complete response (pCR). However, a significant number of TNBC patients develop metastasis and do not survive beyond five years after diagnosis (8). Patients who achieve pCR show a reduced recurrence risk and more prolonged overall survival (OS) (9). The ability to better predict which patients respond to therapy would greatly help clinicians estimate prognosis and guide treatment decisions. In particular, systemic, blood-based predictive biomarkers are of most use in a clinical setting since they are less invasive, easy to collect and return rapid results.

Recent studies have demonstrated that increased stromal tumor-infiltrating lymphocytes (TILs) correlate with better prognosis and response to neoadjuvant therapy in TNBC, while lower TILs are predictive of a worse outcome (10,11). Tumor infiltration by cytotoxic CD8+ T cells and Th1-like CD4+ T cells are correlated with a good prognosis, whereas CD4+ Treg and CD4+ Th2 are linked with unfavorable clinical outcomes (12). These studies demonstrated the importance of the local tumor microenvironment (TME) in modifying tumor response and survival. Peripheral blood immune markers could reflect the local TME composition, and they could also provide clinically relevant information based on circulating soluble factors.

The immune response is influenced by immunologically active cytokines, chemokines, and growth factors, which are produced by a range of cell populations. The relationship between immune cells and immune mediators can influence the formation of an inflammatory TME and lead to a better response to therapy in a variety of cancers (13). In breast cancer, the interaction between tumor cells and Th2-related cytokines can lead to a pro-metastatic phenotype in the TME (14), and higher levels of IL-7 were correlated with worse clinical outcomes in breast cancer patients (15,16). Lower TNF-α concentrations after treatment were associated with a better response to therapy (17,18). In contrast, other studies have suggested that inflammatory cytokines such as IFN-β predict better disease-free survival (19), and higher expression of IL-1β is associated with better overall survival and distant metastasis-free survival (20).

Here we aim to identify systemic immune profiles of pCR (responder) and non-responder (non-pCR) TNBC patients submitted to neoadjuvant chemotherapy by evaluating immune mediators present in plasma before treatment initiation and their correlation with the presence of TILs in biopsies from the same patients. We observed higher levels of inflammatory factors in the blood of patients who obtained pCR. Moreover, these factors alone were predictive of response to therapy, as well as when combined with the percentage of TILs before treatment.

## Results

### 1. Demographics and clinicopathologic characteristics

Our cohort was composed of 39 TNBC patients (18 responder and 21 non-responder). The median age at diagnosis was 41 years old, with no significant difference between groups. The median tumor size at diagnosis was 2.6 cm and was not different between responders or non-responders. However, when the T stage was analyzed individually, we observed significant differences between the groups (p=0.013), with an excess of T3-T4 tumors among non-responders. Distribution of N stage and histological grade distribution was also similar between responder and non-responder patients (Table 1).

**Table 1.**
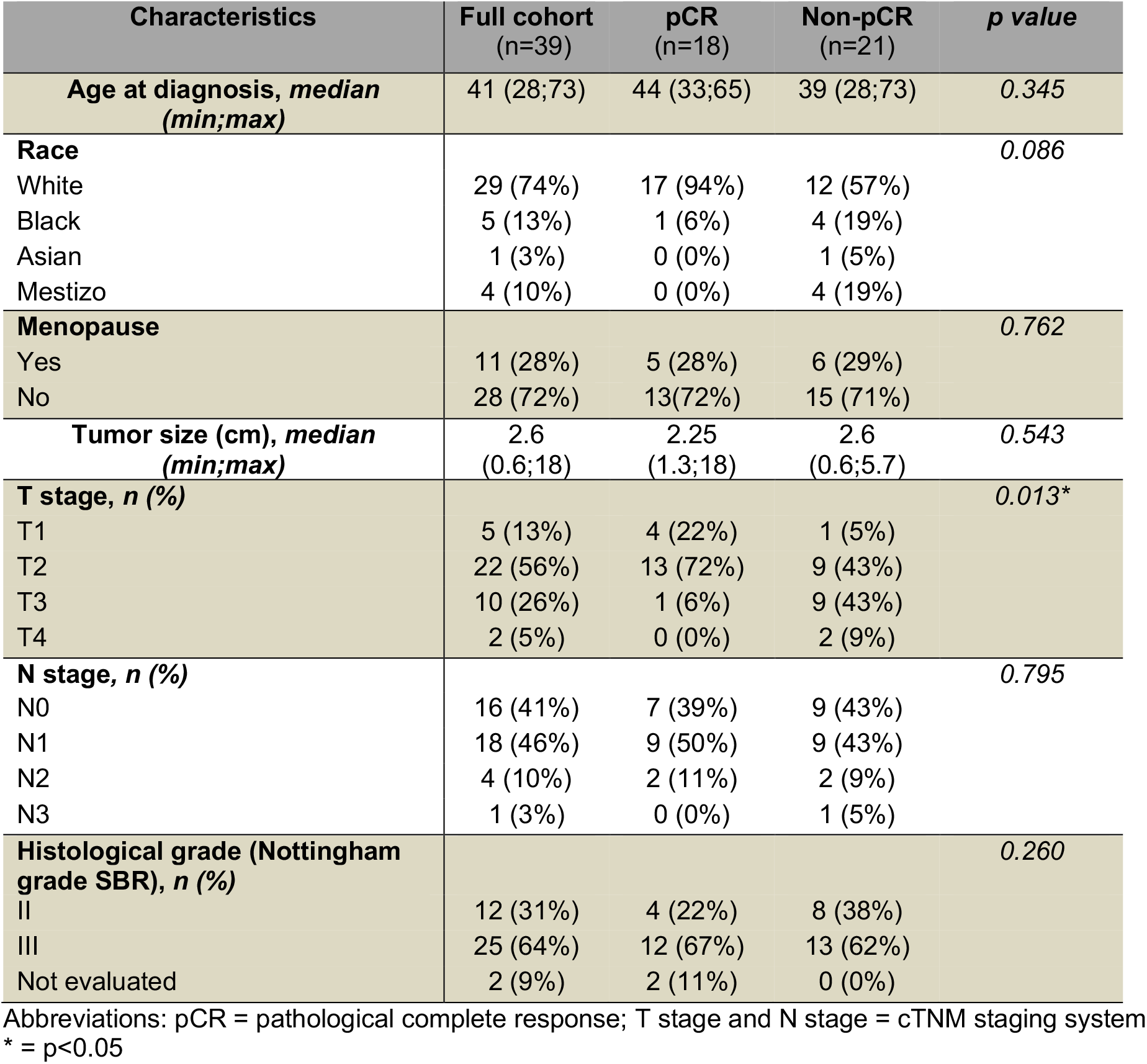
Patient demographics and clinical characteristics.

| Characteristics | Full cohort<br>(n=39) | pCR<br>(n=18) | Non-pCR<br>(n=21) | *p value* |
| --- | --- | --- | --- | --- |
| **Age at diagnosis, median**<br>**(min;max)** | 41 (28;73) | 44 (33;65) | 39 (28;73) | 0.345 |
| **Race** |  |  |  | 0.086 |
| White | 29 (74%) | 17 (94%) | 12 (57%) |  |
| Black | 5 (13%) | 1 (6%) | 4 (19%) |  |
| Asian | 1 (3%) | 0 (0%) | 1 (5%) |  |
| Mestizo | 4 (10%) | 0 (0%) | 4 (19%) |  |
| **Menopause** |  |  |  | 0.762 |
| Yes | 11 (28%) | 5 (28%) | 6 (29%) |  |
| No | 28 (72%) | 13 (72%) | 15 (71%) |  |
| **Tumor size (cm), median**<br>**(min;max)** | 2.6<br>(0.6;18) | 2.25<br>(1.3;18) | 2.6<br>(0.6;5.7) | 0.543 |
| **T stage, n (%)** |  |  |  | 0.013* |
| T1 | 5 (13%) | 4 (22%) | 1 (5%) |  |
| T2 | 22 (56%) | 13 (72%) | 9 (43%) |  |
| T3 | 10 (26%) | 1 (6%) | 9 (43%) |  |
| T4 | 2 (5%) | 0 (0%) | 2 (9%) |  |
| **N stage, n (%)** |  |  |  | 0.795 |
| N0 | 16 (41%) | 7 (39%) | 9 (43%) |  |
| N1 | 18 (46%) | 9 (50%) | 9 (43%) |  |
| N2 | 4 (10%) | 2 (11%) | 2 (9%) |  |
| N3 | 1 (3%) | 0 (0%) | 1 (5%) |  |
| **Histological grade (Nottingham**<br>**grade SBR), n (%)** |  |  |  | 0.260 |
| II | 12 (31%) | 4 (22%) | 8 (38%) |  |
| III | 25 (64%) | 12 (67%) | 13 (62%) |  |
| Not evaluated | 2 (9%) | 2 (11%) | 0 (0%) |  |

Abbreviations: pCR = pathological complete response; T stage and N stage = cTNM staging system
\* = $p<0.05$

### 2. Responder patients display higher levels of plasma immune mediators when compared with non-responders

To investigate the possible association between circulating cytokines, chemokines and growth factors with a response to therapy as determined by pCR, we performed multiplex bead assays of 27 immune mediators in patient plasma collected before patients began treatment. We observed higher levels of FGF-basic and VEGF (growth factors), as well as GM-CSF and IL-2 (secreted by activated immune cells) in responding patients (Fig 1A). The same was observed for the Th2 associated cytokine, IL-5 (Fig 1B). IL-13 and IL-15 displayed a slight increase in the responder group with *p* values less than 0.10. Concentrations of TNF-α, IL-7 and IFN-γ in the responder group had slightly elevated median levels in the responder group, but with *p* values >0.10. (Supplementary Fig 1). These results demonstrate that TNBC patients that will go on to respond to therapy with pCR display higher levels of some circulating cytokines indicative of an activated immune system.

**Figure 1.**
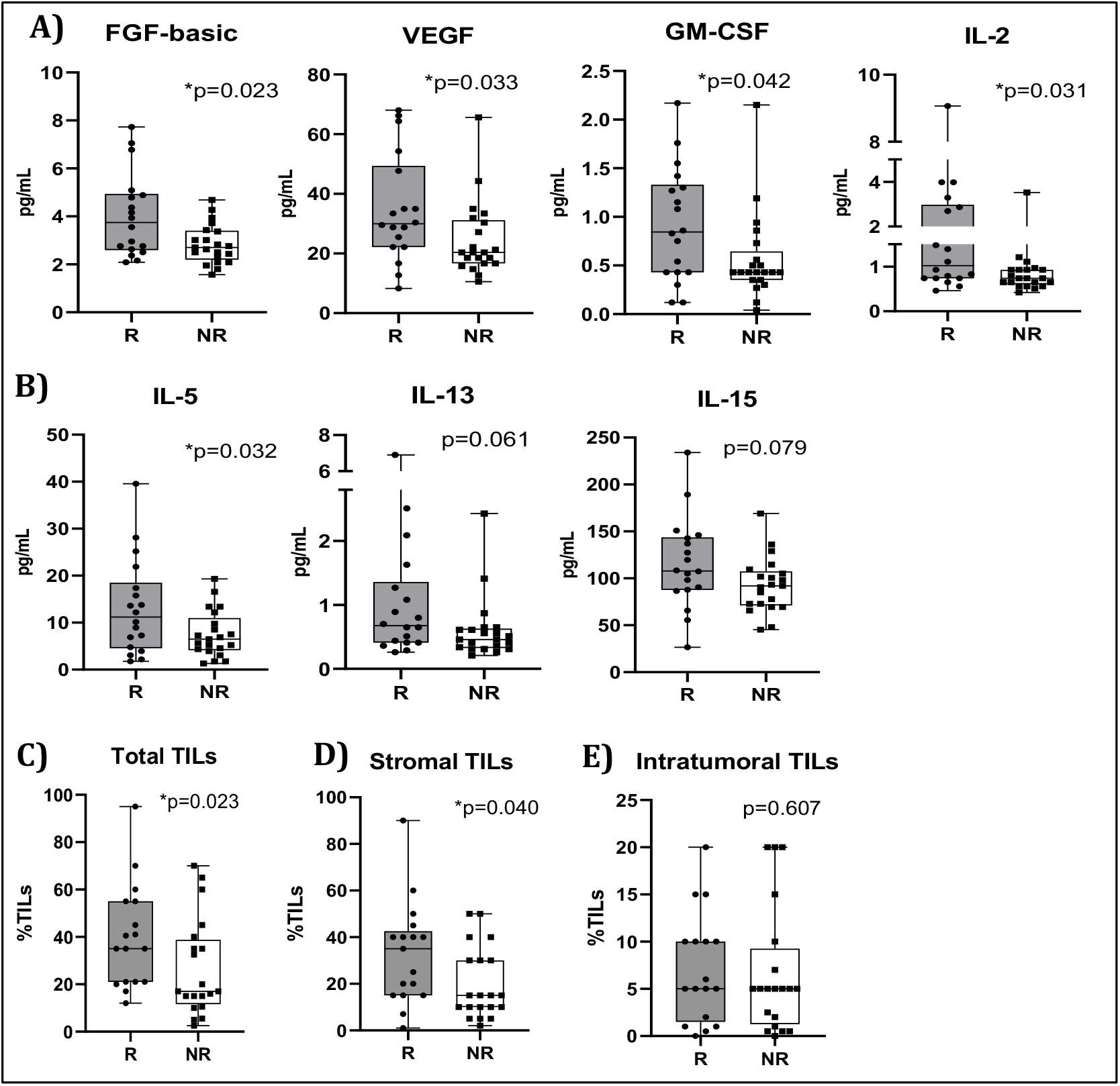
Blood from TNBC patients submitted to neoadjuvant chemotherapy was collected at the time of diagnosis. Immune mediators were quantified in the plasma of patients who obtained complete pathological response (responders – R (n=18) or not (non-responders – NR) (n=21) using Bio-Plex assay. (A) FGF-b, VEGF, GM-CSF, and IL-2 quantification. (B) IL-5, IL-13, and IL-15 quantification. TIL counts were analyzed in H&E stained slides obtained at the time of diagnosis TNBC of patients who obtained complete pathological response after neoadjuvant chemotherapy (responders – R) (n=17) or not (non-responders – NR) (n=20). (C) Total TILs (Stromal + Intratumoral TILs). (D) Stromal TILs. (E) Intratumoral TILs. All statistical analyses were performed with the Mann-Whitney test (FGF-b, VEGF, GM-CSF, IL-2, Total TILs, Stromal TILs and Intratumoral TILs) or the Student’s t test (IL-5 and IL-15) (*p<0,05). Immune mediators are expressed in pg/ml.

### 3. Systemic inflammatory cytokines are positively correlated with the levels of TILs in pCR responder patients

It is widely recognized that the presence of high TIL levels is predictive of response to neoadjuvant chemotherapy and correlate with favorable outcome in TNBC patients (23). Moreover, given the increase in several systemic cytokines in pCR responder patients, we performed an analysis to determine if systemic cytokine levels were correlated with percentage of TILs in the responders and non-responders. Thus, we first evaluated TILs in the stromal and intratumoral compartments using H&E stained slides to assess their association with response to treatment (Supplementary Fig 4). Responders displayed higher levels of both total TILs (percentage of stromal + intratumoral TILs) and stromal TILs when compared with non-responders, as expected (Fig 1C and 1D). Intratumoral TILs were equivalent between the two groups (Fig 1E). These results suggest that a more immunogenic TME is associated with therapy response in TNBC patients.

Next, the levels of circulating cytokines were plotted vs. TILs percentage in each patient. We observed that levels of IFN-γ were positively correlated with total TILs in responders (Fig 2A), and intratumoral TILs were positively correlated with IL-7 in the same group (Fig 2B). In contrast, TNF-α levels were negatively correlated with stromal TILs in non-responder patients, while no significant correlation was seen in responder patients (Fig 2C).

**Figure 2.**
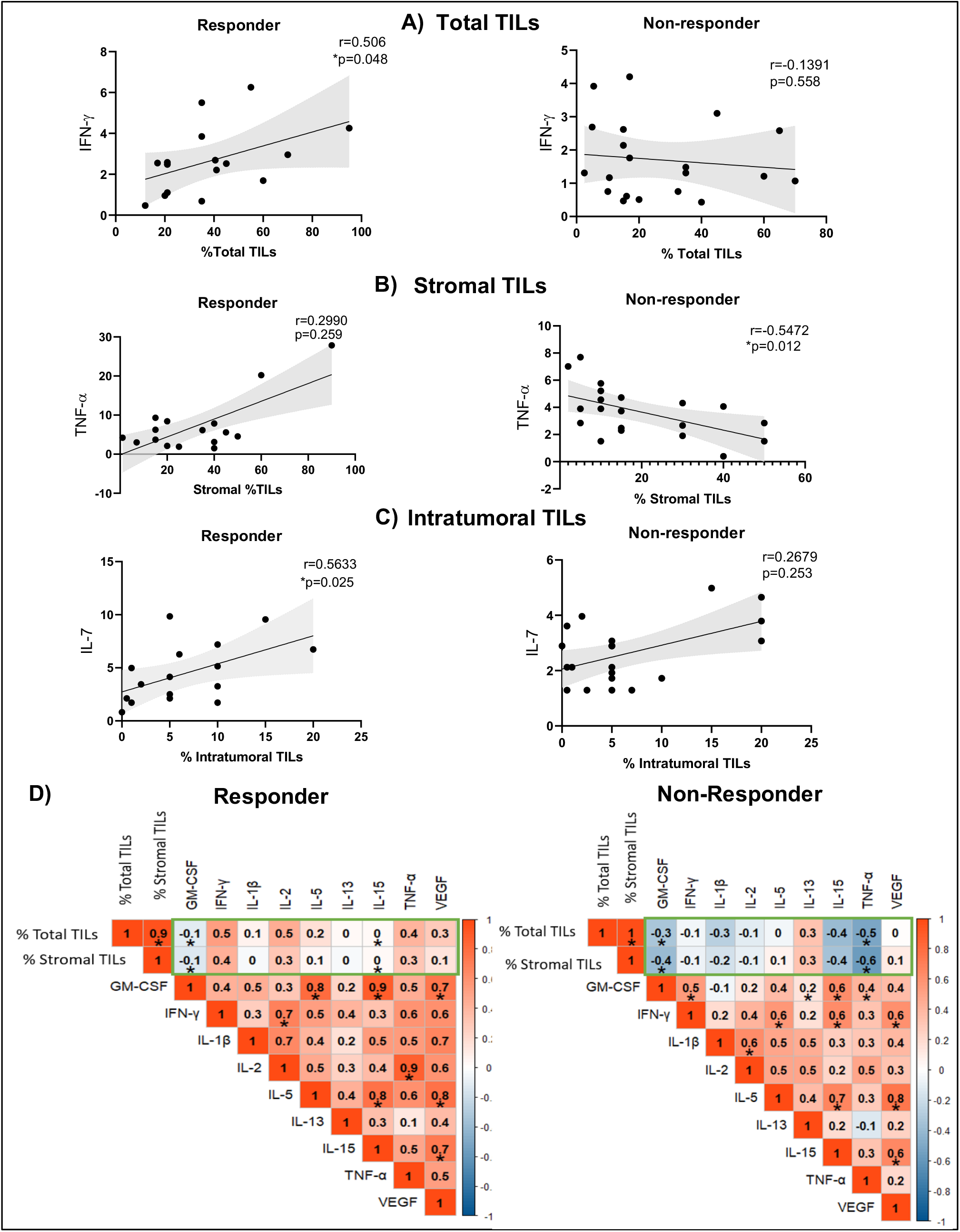
Correlation between TILs and cytokines in patients who obtained complete pathological response after neoadjuvant chemotherapy classified as Responders (n=16) or Non-responders (n=20). (A) Correlation between Total TILs and plasma IFN-γ. (B) Correlation between Stromal TILs and plasma TNF-α. (C) Correlation between Intratumoral TILs and plasma IL-7. (D) Correlation matrix between cytokines and TILs in responder and non-responder group. All statistical analyses were performed with the Spearman’s correlation Test (*p<0.05). Immune mediators are expressed in pg/ml.

To obtain an overall view of the immunoregulatory network between responders and non-responder patients we generated correlation matrices of key soluble immune factors, total TILs and stromal TILs in the responder and non-responder groups. We observed a stronger positive correlation between several inflammatory cytokines and TIL levels, as well as between cytokines in the responder group as compared to the non-responders (Fig 2D). Correlation matrices were also generated with all 27 cytokines and TILs (total, stromal and intratumoral) in both groups (Supplementary Fig 2), which shows a trend similar to that seen with the smaller group of cytokines, with particularly strong positive correlation between soluble immune factors and percentage of TILs, as compared to a negative or weaker correlation for the non-responder group.

Together, these data support a stronger association between inflammatory circulating cytokines and tumor-infiltrating lymphocytes in TNBC patients that will respond to therapy with pCR.

### 4. Systemic soluble cytokines and TILs predict clinical outcome in TNBC

Given the association of systemic cytokines with response to treatment and their correlation with TILs, our next step was to evaluate whether these molecules were associated with clinical outcome. Using logistic regression, we observed that the cytokines FGF-basic, IL-2, IL-5, IL-7, TNF-α, VEGF, IFN-γ and GM-CSF were independent predictors of pCR (Table 2).

**Table 2.** Univariable and Multivariable Analyses of Cytokines and TILs for prediction of pCR.

|  |  |  |  | MULTIVARIABLE ANALYSES |  |  |  |  |  |
| --- | --- | --- | --- | --- | --- | --- | --- | --- | --- |
|  | UNIVARIABLE ANALYSES |  |  | Total TILS |  |  | Stromal TILS |  |  |
| CYTOKINE | OR | 95% CI | P Value | OR | 95% CI | P Value | OR | 95% CI | P Value |
| **FGF-b** | 2.16 | 1.22 to 4.63 | **.006** | 2.22 | 1.20 to 5.00 | **.027** | 2.24 | 1.20 to 5.04 | **.025** |
| **IL-5** | 1.11 | 1.01 to 1.25 | **.024** | 1.15 | 1.02 to 1.33 | **.037** | 1.14 | 1.02 to 1.33 | **.039** |
| **GM-CSF** | 4.02 | 1.10 to 20.00 | **.035** | 6.73 | 1.53 to 47.66 | **.026** | 7.07 | 1.58 to 52.9 | **.025** |
| **TNF-α** | 1.26 | 1.02 to 1.76 | **.027** | 1.38 | 1.02 to 2.06 | **.021** | 1.41 | 1.03 to 2.12 | .067 |
| **IL-2** | 2.38 | 1.17 to 7.47 | **.009** | 2.50 | 1.14 to 8.53 | .059 | 2.57 | 1.13 to 8.76 | .060 |
| **IL-7** | 1.44 | 1.03 to 2.20 | **.031** | 1.54 | 1.02 to 2.65 | .068 | 1.52 | 1.03 to 2.53 | .060 |
| **VEGF** | 1.05 | 1.00 to 1.11 | **.035** | 1.06 | 1.01 to 1.13 | .078 | 1.06 | 1.00 to 1.13 | .053 |
| **IFN-γ** | 1.61 | 1.00 to 2.86 | **.048** | 1.59 | 0.94 to 2.99 | NS | 1.55 | 0.911 to 2.91 | NS |
| **IL-15** | 1.02 | 1.00 to 1.04 | .068 | 1.02 | 1.00 to 1.05 | .057 | 1.02 | 1.00 to 1.05 | .061 |
| **IL-13** | 2.50 | 1.00 to 11.13 | NS | 2.55 | 0.95 to 13.91 | NS | 2.43 | 0.955 to 13.6 | NS |

Abbreviations: OR = odds ratio; 95% CI = confidence interval; NS = non statistically significant

Once we identified that individual cytokines predicted pCR, we performed a multivariable logistic regression where total and stromal TILs were identified as significant co-variants. The multivariate analysis adjusted for TILs showed an increased capacity of cytokines to predict pCR, like FGF-basic, IL-2, IL-5, IL-7, TNF-α, VEGF and GM-CSF (Table 2). In particular, GM-CSF alone returned an OR of 4.02 (1.10 to 20.00), p =0.035, while together with the percentage of stromal TILs they returned an OR of 7.07 (1.58-52.9), p=0.025. To investigate the sensitivity and specificity of soluble cytokines as a predictor of pCR we performed a series of univariate and multivariate ROC curves, which returned AUC from 0.64 to 0.71 for cytokines alone (Fig 3). AUC analysis for stromal and total TILs alone was 0.70 and 0.72, respectively. Thus, several of the systemic cytokines alone with AUC >0.70 (IL-2, VEGF, and GM-CSF) were similar to TILs in their ability to identify responder vs. non-responder individuals. When combining cytokines with TIL levels, all returned increased AUC (range of 0.71 - 0.82) (Fig 3). TILs alone showed a lower capacity to predict response compared to the other analyses (Supplementary Table 1).

**Figure 3.**
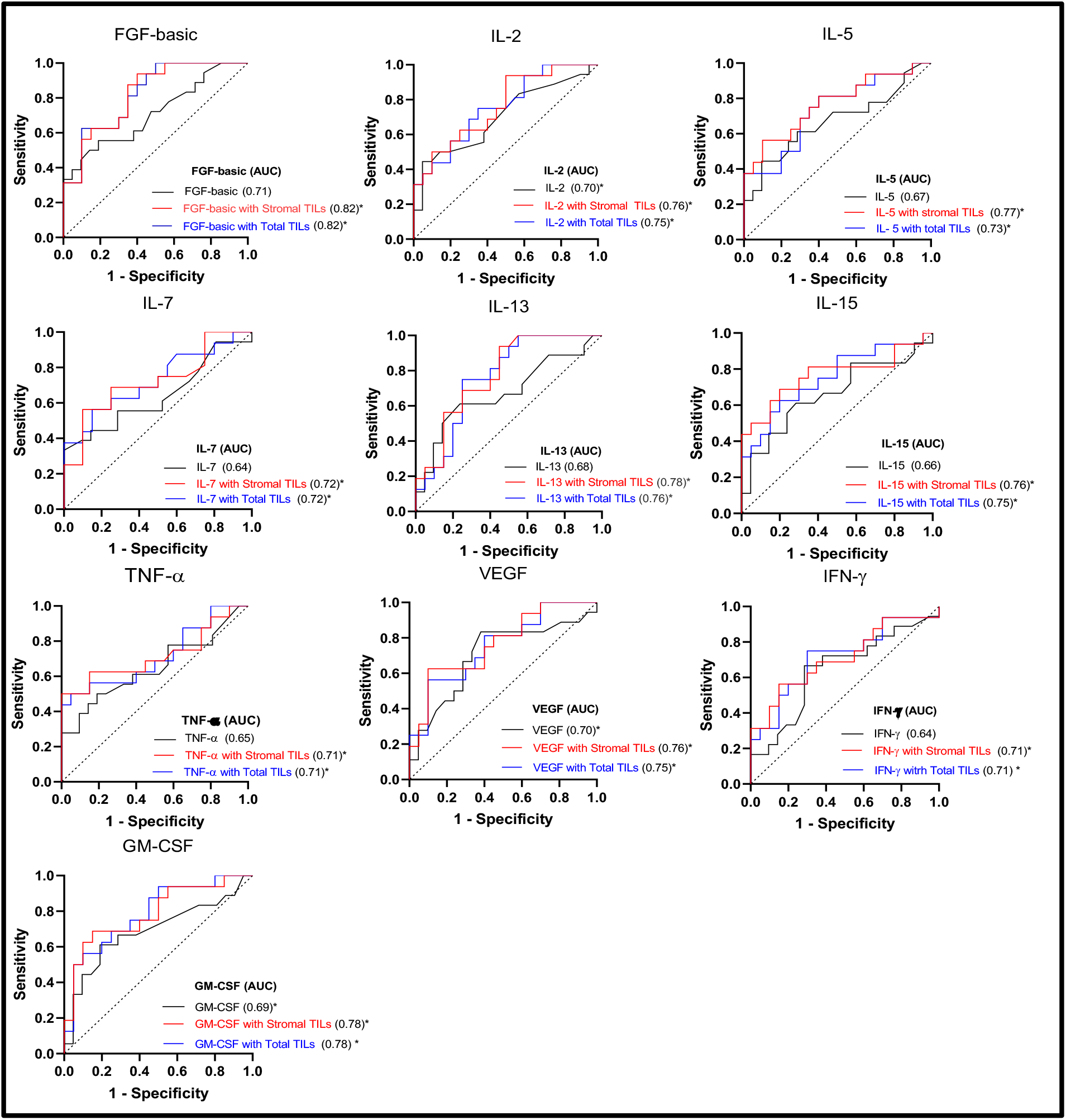
The accuracy of cytokine as biomarkers was evaluated using receiver operating characteristic (ROC) curves, and the Area Under the Curve (AUC) was calculated for each cytokine (black) (univariable analysis) and in combination with Stromal TILs (red) or combination with Total TILs (blue) (multivariable analysis) (*p<0.05).

Analyses considering cytokines with TILs and histological grade were also performed. Similarly, we performed analyses adjusted for histological grade only, with the addition of total or stromal TILs (Supplementary Table 2). However, in both scenarios, no great improvement was seen over cytokine and TILs alone (Supplementary Fig 3).

Analyses in combination with intratumoral TILs showed improvement in performance for some cytokines, but several lost significances (Supplementary Table 3).

These results suggest that TNBC patients with increased levels of specific circulating cytokines associated with higher levels of total or stromal TILs before treatment have a better chance of achieving a pathological complete response.

### 5. In silico analysis reveals better survival in TNBC patients with high tumor expression of IL-7, IL-15, TNF-α, and IFN-γ

Given our findings showing the plasma cytokines’ value to predict pCR, we used a bioinformatics approach, based on the TCGA database, to validate our findings using transcriptomic data. We observed that TNBC patients with higher tumor mRNA expression of IL-7, IL-15, TNF-α (TNFSF2) and IFN-γ had longer Relapse free-Survival (RFS) (Fig 4A), which is in line with our findings that plasmatic levels of IL-7, TNF-α, and IFN-γ are predictive of pCR (Table 2). Moreover, both IL-7 and IFN-γ were also positively correlated with TILs in responding patients (Figure 2), and TNF-α negatively correlated with TILs in non-responders. In contrast, although responders had higher plasmatic levels of FGF-basic (FGF1), IL-13, VEGF (VEGFA) and GM-CSF (CSF2), the expression of the corresponding genes in the tumor was associated with shorter RFS (Fig 4B).

**Figure 4.**
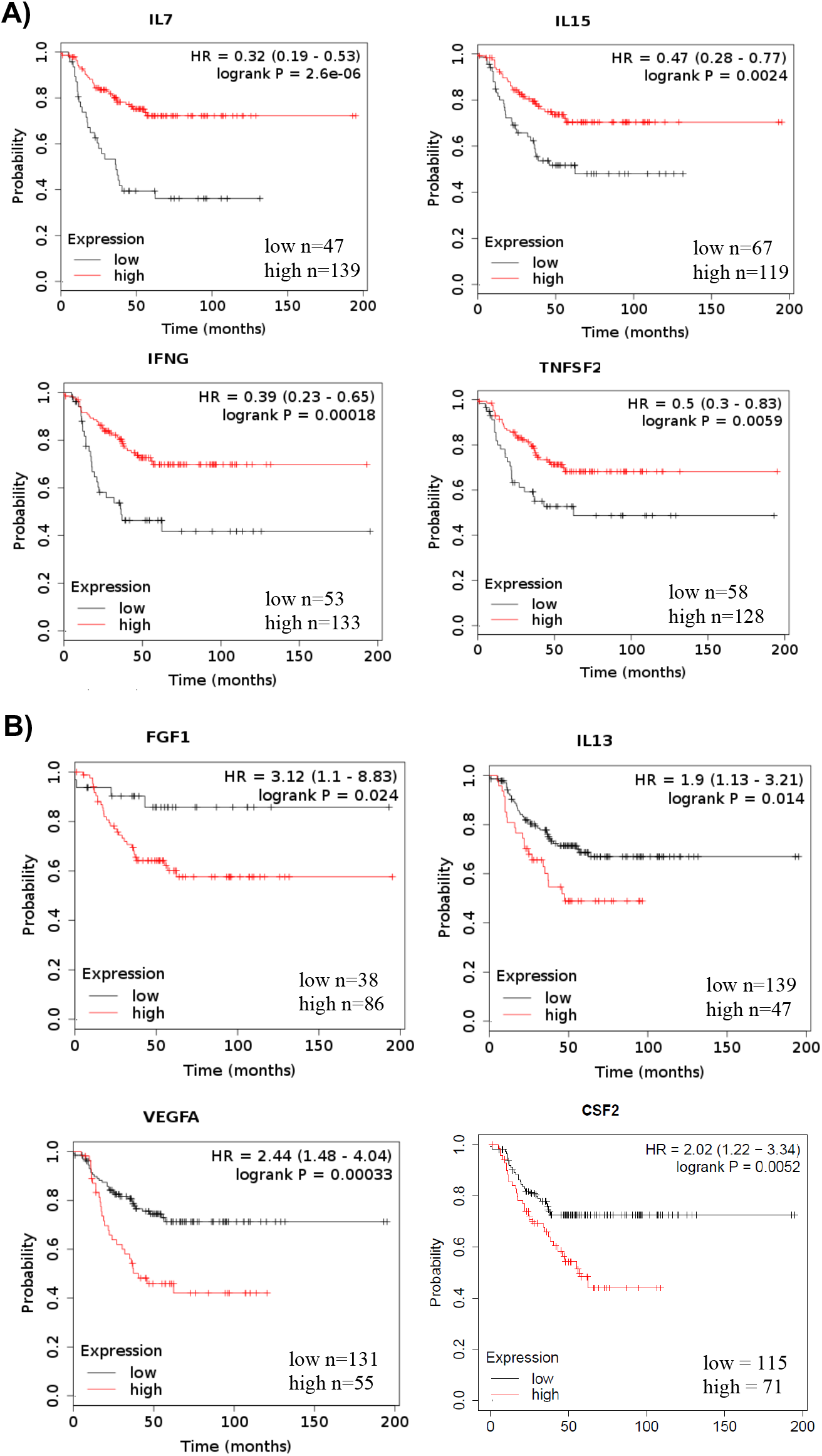
Relapse-free survival (RFS) analysis comparing TNBC patients with high and low expression of inflammatory cytokines genes. These analyses were performed using TCGA data available in KM Plotter. (A) RFS curves according to *IL-7, IL-15, TNFSF2* (TNF-α) and *IFNG* (IFN-γ) gene expression in TNBC tumor samples. (B) RFS curves according to *FGF1* (FGF-basic), *IL-13, VEGFA* (VEGF) and *CSF2 (*GM-CSF) gene expression in TNBC tumor samples. Survival curves were compared by log-rank test.

## Discussion

Cytokines and chemokines are essential for systemic and tumor microenvironment activation or inhibition of immune responses (24), and numerous studies have demonstrated that higher levels of TILs in the TME are often associated with better patient outcomes (25). In contrast, tumor cells can produce cytokines capable of modulating the immune system to create an immunosuppressed microenvironment favorable to tumor growth (26). These molecules can be reflected in the circulation providing a window into potentially important immunoregulatory networks with clinical relevance. For this reason, cytokines present in plasma before a patient begins treatment may reflect the current immune system status, as well as, provide potential biomarkers to predict treatment response and clinical outcome.

IL-2 is a key cytokine responsible for lymphocyte proliferation, maturation and activation (27). In our study, we saw increased levels of IL-2 in responding patients compared to non-responders (Fig 1). Moreover, our correlation matrices showed an association between proinflammatory cytokines and TILs in responders when compared to non-responders (Fig 2). A Japanese study with breast cancer patients noted that higher TIL counts were correlated with increased levels of IL-2 and IL-1β and low levels of eotaxin (28). This finding suggests that responding TNBC patients may have a more activated immune profile than non-responding ones. Interestingly, our results also showed that TNBC responding patients had higher levels of IL-5 and IL-13 (p = 0.06) in the plasma (Fig 1), while previous studies have shown high serum levels of Th2 cytokines associated with worse prognosis in patients with other breast cancer subtypes (14). Other studies have indicated that tumors are capable of inducing systemic inflammation, hematopoietic tissue reprogramming and promote myeloid cell elevation by secreting cytokines and growth factors that enable disease progression (26,29).

In our study we observed that responding patients had higher plasma levels of FGF-basic, VEGF and GM-CSF, which act as growth factors for some cell populations and promote vessel formation (Fig 1). However, previous studies have reported contradictory results regarding their role in cancer. Ghirelli et al. (2015) demonstrated that breast cancer cell-derived GM-CSF was capable of promoting a regulatory response through Th2 phenotype modulation leading to an aggressive disease subtype (30). In another study, breast cancer cell-derived FGF was seen to promote osteoclast differentiation *in vitro* and was associated with bone metastasis in murine models (31). Higher intratumoral levels of VEGF was observed in TNBC patients when compared with other breast cancer subtypes (32). All these findings may contribute to improve our understanding of how the immune system modulates the response to chemotherapy in TNBC patients, since higher plasma levels of pro-inflammatory cytokines, growth factors and Th2 cytokines were detected in patients that achieved pCR compared to the non-responder group. The higher levels of Th2 could act as a counterbalance to an amplified systemic inflammation creating a homeostasis in responder patients.

Recent evidence suggests that the high levels of stromal TILs in TNBC biopsies is a predictive factor of pCR to neoadjuvant chemotherapy (11,33,34). In our study, we observed higher levels of total and stromal TILs associated with treatment response and pCR (Fig 1). In addition, we observed a positive correlation between pro-inflammatory cytokines (IFN-γ and IL-7) and both total and intratumoral TILs in responding patients (Fig 2A,C) and a negative correlation between TNF-α and stromal TILs in non-responding patients (Fig 2B). Importantly, we observed an overall pattern in responder patients of positive correlations between TILs and inflammatory cytokines, while non-responder patients displayed negative correlations with TILs for the same cytokines (Fig 2D). TILs and cytokines were correlated in other studies involving breast cancer patients, revealing an association between higher levels of pro-inflammatory cytokines like IP-10 and RANTES and TILs, indicating that monitoring circulating cytokines could help to characterize the tumor microenvironment (14,35) and serve as predictive biomarkers.

Our results showed an association between the cytokines IL-2, IL-7, TNF-α and IFN-γ and responder patients as determined by increased levels, correlation with TILs, or for their prediction of pCR (Figs 1,2 and Table 2). These results were reinforced by *in silico* analysis in which we found higher mRNA levels of IL-7, TNF-α and IFN-γ associated with better relapse free survival (Fig 4). In contrast, higher levels of IL-13, VEGF, FGF-b and GM-CSF messenger RNA were associated with worse RFS, while the same cytokines were increased in the plasma of patents with pCR (Fig 1). IL-2 mRNA levels did not show any significant differences in RFS. These contradictory findings clearly suggest these cytokines have different roles in the local and systemic immune responses against breast cancer and this can be due to several factors such as T cell activation and suppression, differential tissue homing of immune cell subpopulations, bone marrow reprograming and mobilization of myeloid cells, all processes that are directly associated with clinical outcome. It will be interesting to follow the patients included in this cohort and investigate if the levels of cytokines and growth factors in the blood will also correlate with long-term survival.

In conclusion, our study demonstrates that circulating proinflammatory cytokines are associated with pCR in TNBC patients treated with neoadjuvant chemotherapy and may act as a surrogate marker of high TILs or together with TILs to better predict pCR and survival.

## Methods

### Patients and classification

We included 39 immunocompetent women diagnosed with non-metastatic TNBC that underwent neoadjuvant chemotherapy based on anthracycline, carboplatin and taxanes followed by surgery and radiation therapy. This study was approved by the local Ethics Committee. A written informed consent was obtained from each patient at the time of diagnosis and before the initiation of any procedure. All patients were recruited at A.C.Camargo Cancer Center (ACCCC, São Paulo, Brazil). Hematoxylin & Eosin (H&E) - stained slides were obtained from formalin fixed and paraffin embedded (FFPE) tissue blocks representative of core needle breast biopsies and stored at the Department of Anatomic Pathology of ACCCC. Tumor staging (TNM) was recovered from medical charts. The clinical, pathological and immunophenotypic data of the patients were obtained from the available electronic clinical files. Blood samples were collected before the initiation of any therapy and plasma immune mediator levels were evaluated from all patients.

Class of pathologic response to neoadjuvant treatment was determined according to the Residual Cancer Burden (RCB) protocol at time of surgery (21). Pathologic complete response (pCR) corresponds to RCB = 0 and it is defined by the absence of invasive carcinoma in the breast and axillary lymph nodes. These patients are considered “responders” in this study. Patients with a RCB > 0 did not have pCR, and were classified as a group (RCB = 1, 2, or 3) as “non-responders” in this study. Relapse-free survival (RFS) was defined as the time from surgery to the first occurrence of local or distant disease recurrence (22).

### Assessment of Tumor Infiltrating Lymphocytes

Levels of TILs were evaluated in 37 samples. From each selected patient histopathological determination of TILs was performed in H&E stained slides according to the International TILs Working Group criteria (12). Intratumoral and stromal TILs were quantified as the percentage of tumor area occupied by mononuclear inflammatory cells (lymphocytes and plasma cells) by a breast pathologist blinded to levels of circulating immune mediators and treatment outcome (Supplementary Figure 4).

### Plasma immune mediator quantification

A total of 27 molecules including cytokines, chemokines and growth factors (FGF-basic, Eotaxina, G-CSF, GM-CSF, IFN-γ, IL-1β, IL-1ra, IL-2, IL-4, IL-5, IL-6, IL-7, IL-8, IL-9, IL-10, IL-12p70, IL-13, IL-15, IL-17a, IP-10, MCP-1 (MCAF), MIP-1α, MIP-1β, PDGF-B, Rantes, TNF-α and VEGF) were measured in the plasma samples using a soluble factor multiplex bead assay (Bio-plex Pro Human Cytokine Grp I panel 27-plex, BioRad Inc, USA). Samples were processed by the core facility at the René Rachou Institute – Fiocruz (Minas Gerais, Brazil) using the Bio-Plex® 200 system (BioRad Inc., USA) and all data was acquired using the Bio-Plex Manager software. Values are reported as pg/ml using standard curves provided by the manufacture. Plasma samples were prepared from peripheral blood collected in EDTA tubes (Becton Dickenson, USA) by centrifugation at 400g for 15 mins to remove the plasma and then frozen at −80^°^C within 4 hours of collection.

### Bioinformatics analysis

To further investigate our findings using an expanded number of patients, we analyzed TCGA and GTEx data available in web-based databanks hosting cancer-related molecular characteristics and clinical data. The correlation between (*IL7, IL15, TNFSF2, IFNG, CSF1, FGF1, IL13, VEGFA, IL-17A, IL2* and *IL5*) gene expressions and RFS in TNBC samples compared with normal samples were analyzed by Kaplan-Meier estimator (http://kmplot.com/analysis/) (19). The cutoff for high and low expression of the genes was automatically calculated by KM plotter algorithm. The Cox semiparametric proportional hazards model (Cox, 1972) was fitted to assess which variables would be associated with the endpoints. The assumption of proportional hazards was based on the Schoenfeld residuals.

### Statistical Analysis

The baseline patient characteristics are expressed as absolute and relative frequencies for qualitative variables and as the median, minimum and maximum for quantitative variables. The association between qualitative variables was evaluated by chi-squared test or Fisher’s exact test, as appropriate. To compare immune mediators and TILs between responders and non-responders we used Mann-Whitney or Student’s t test, depending on the data distribution after testing for normality. The correlation between immune mediator levels and TILs was performed using the Spearman’s correlation. In order to evaluate the effect of each immune mediator in the outcome pCR (responders) a simple logistic regression model was fitted to the dataset to estimate the odds ratios (OR) and its 95% confidence intervals (95%CI). In addition, we fitted a multiple logistic regression model considering immune mediators, TILs and histological grade as independent variables. For model performance, its capacity to discriminate between events and nonevents was assessed through receiver operating curves (ROC) curves and the C-statistic, which represents the area under the ROC curve (AUC). The goodness of fit was carried out through of the Hosmer-Lemeshow test.

The significance level of the tests was fixed at 0.05. The statistical analyzes were performed using R software version 3.5 (R Development Core Team) and GraphPad Prism 8.4 software.

## Data Availability

The datasets generated during the current study are available from the corresponding author on reasonable request.

## Acknowledgements

This work was funded with support from CNPq, PRONON, and INCT-INCITO/CNPq. KJG, DMC, ATC are CNPq research fellows. FAPESP, CAPES and CNPq for graduate student fellowships.

---

**Supplementary Table 1.** Univariable Analyses of TILs levels for prediction of pCR.

|  | UNIVARIABLE ANALYSES |  |  |
| --- | --- | --- | --- |
|  | TILs (%) |  |  |
|  | OR | 95%CI | p-value |
| **Total TILs** | 1.03 | 0.99 to 1.07 | NS |
| **Stromal TILs** | 1.04 | 1.003 to 1.09 | .035 |
| **Intratumoral TILs** | 1.01 | 0.91 to 1.12 | NS |

Abbreviations: OR = odds ratio; 95% CI = confidence interval; NS = non statistically significant

**Supplementary Table 2.**
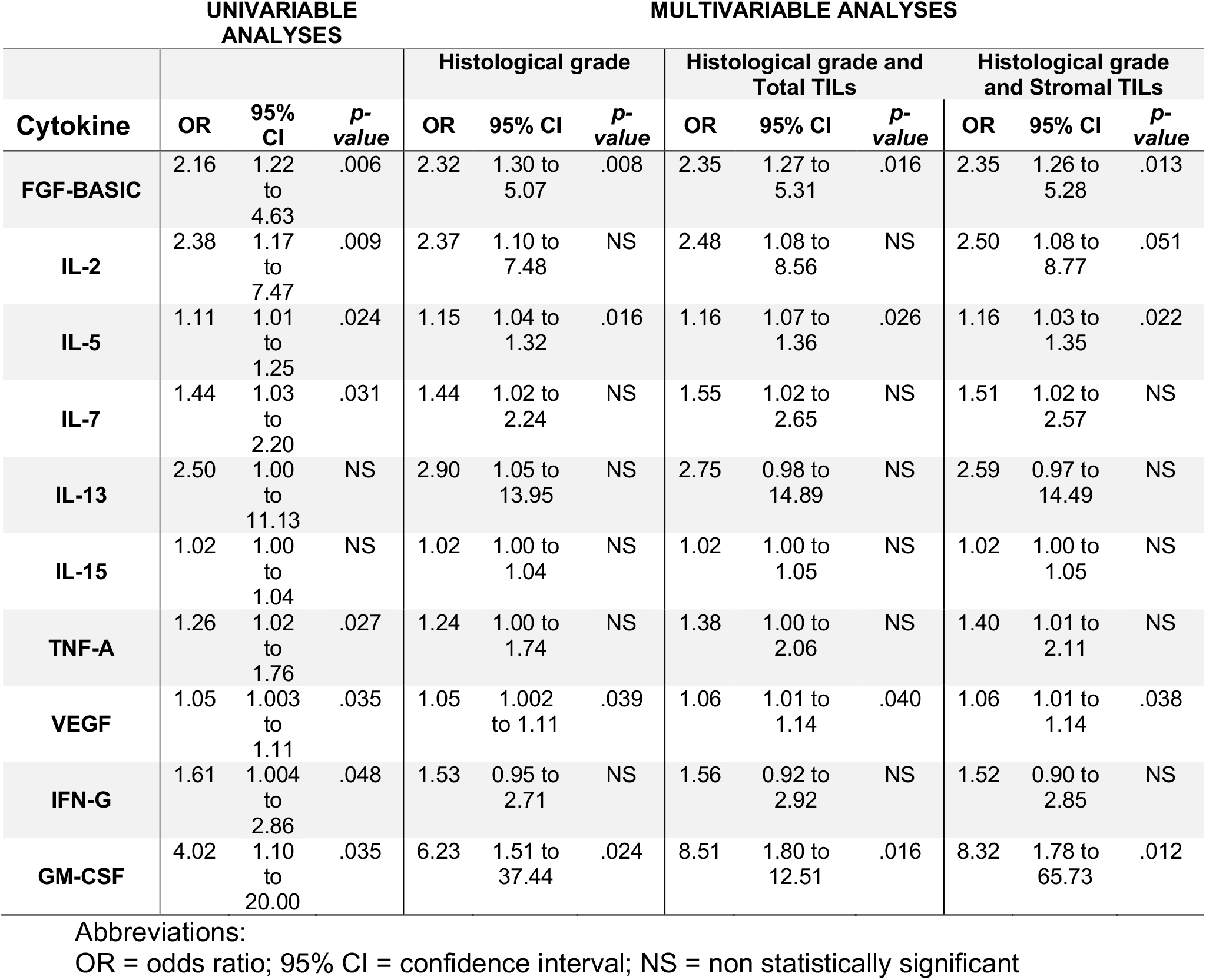
Univariable and Multivariable Analyses of Cytokines, Histological grade and TILs for prediction of pCR.

**Supplementary Table 3.**
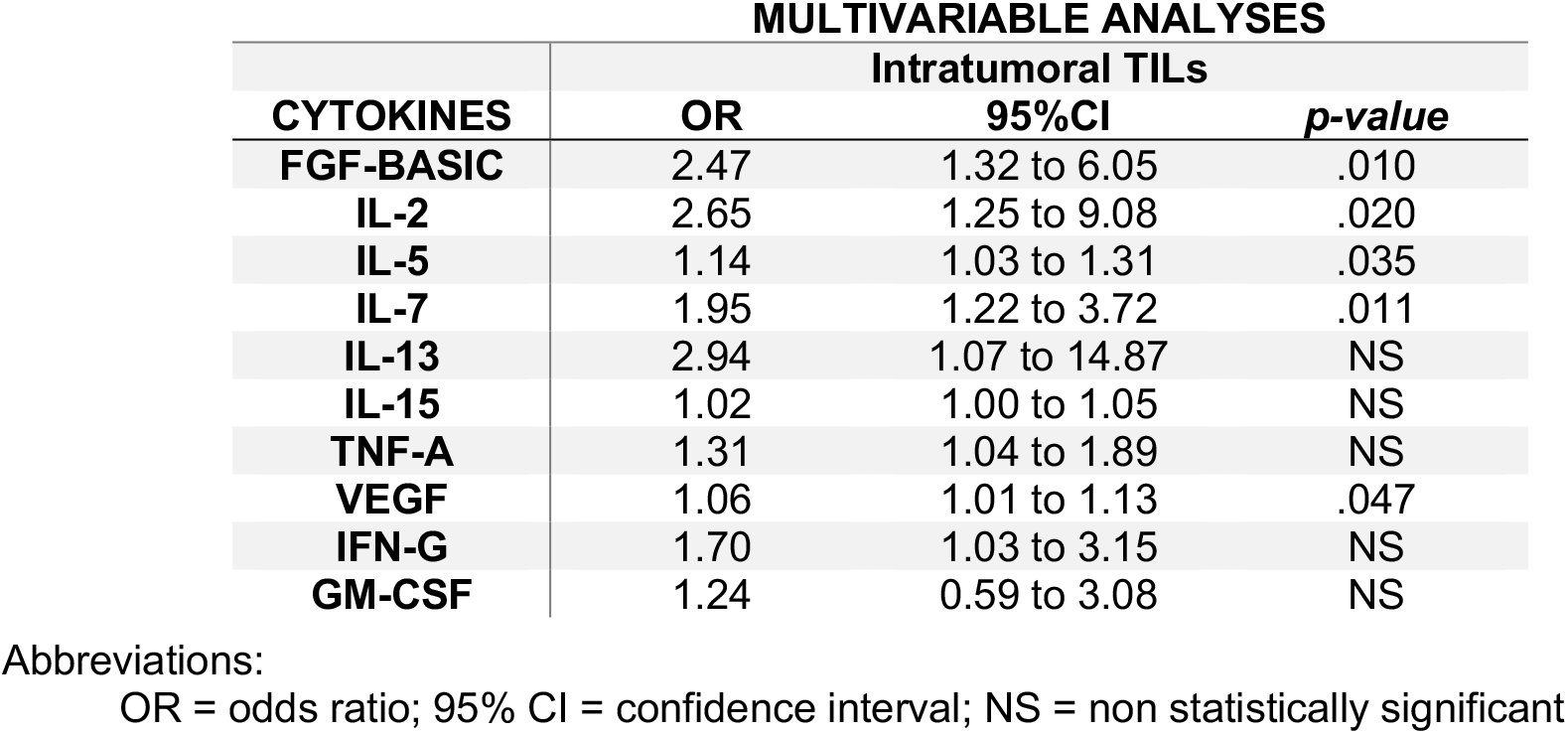
Multivariable Analyses of Cytokines with Intratumoral TILs for prediction of pCR.

## Supplementary Figures

**Supplementary Figure 1.**
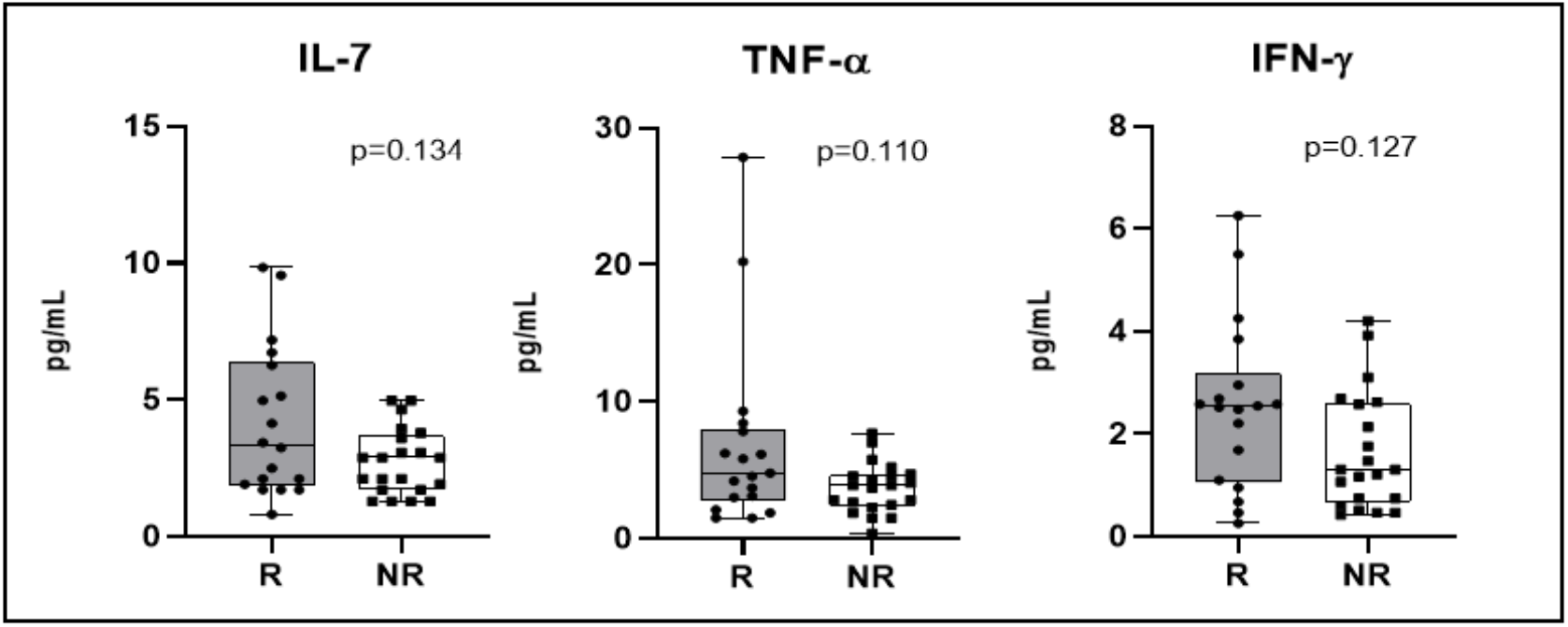
Cytokines were quantified in the plasma (collected at the time of diagnosis) of patients who obtained complete pathological response after neoadjuvant chemotherapy (responders – R) (n=18) or not (non-responders – NR) (n=21) using Bioplex assay. IL-7, TNF-α and IFN-γ quantification. All statistical analyses were performed with the Mann-Whitney test (*p<0,05).

**Supplementary Figure 2.**
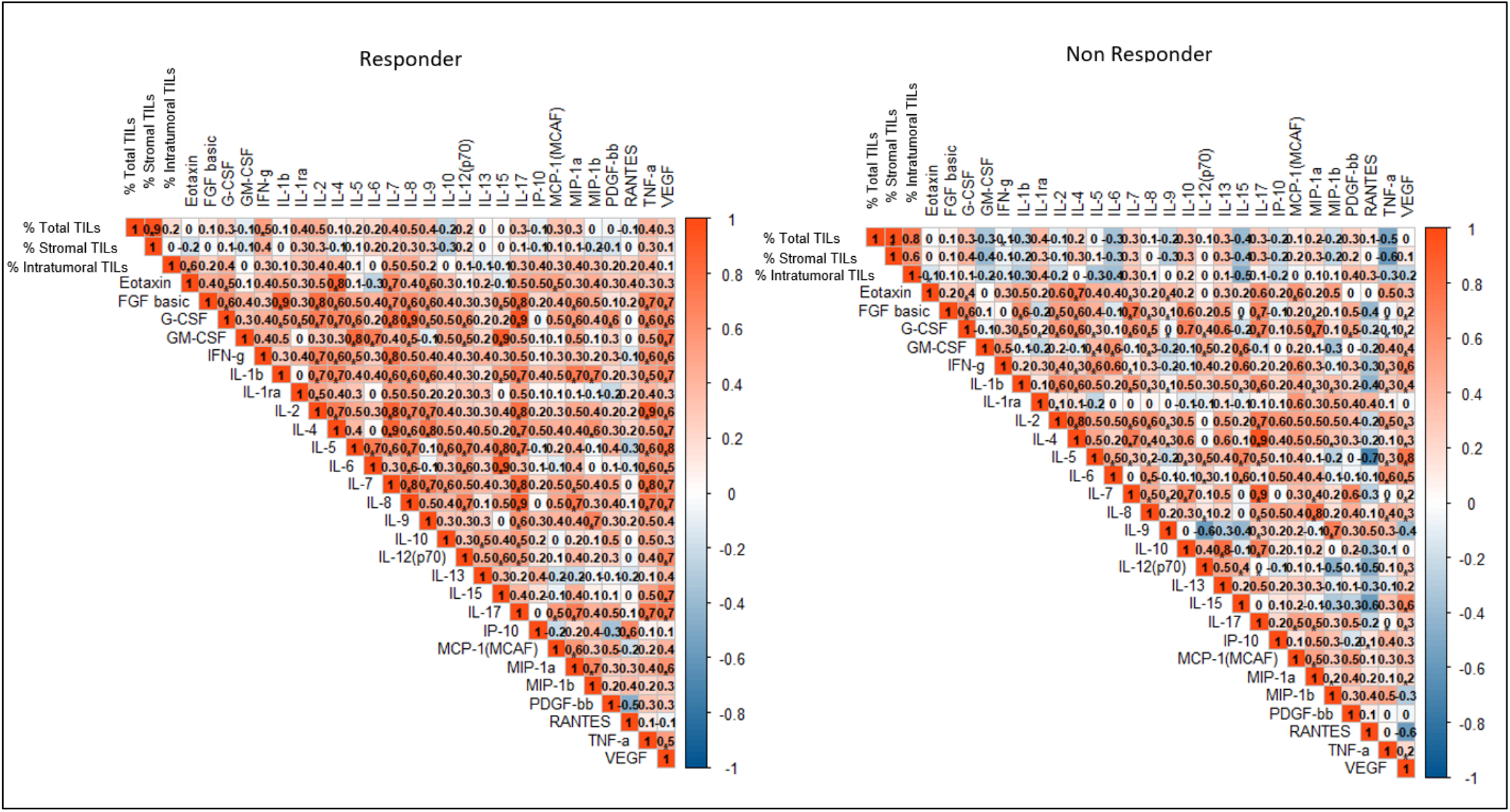
Correlation matrix of all 27 cytokines and TILs (Total, Stromal and Intratumoral) in the responder and non-responder groups. All statistical analyses were performed with the Spearman’s correlation Test (*p<0.05).

**Supplementary Figure 3.**
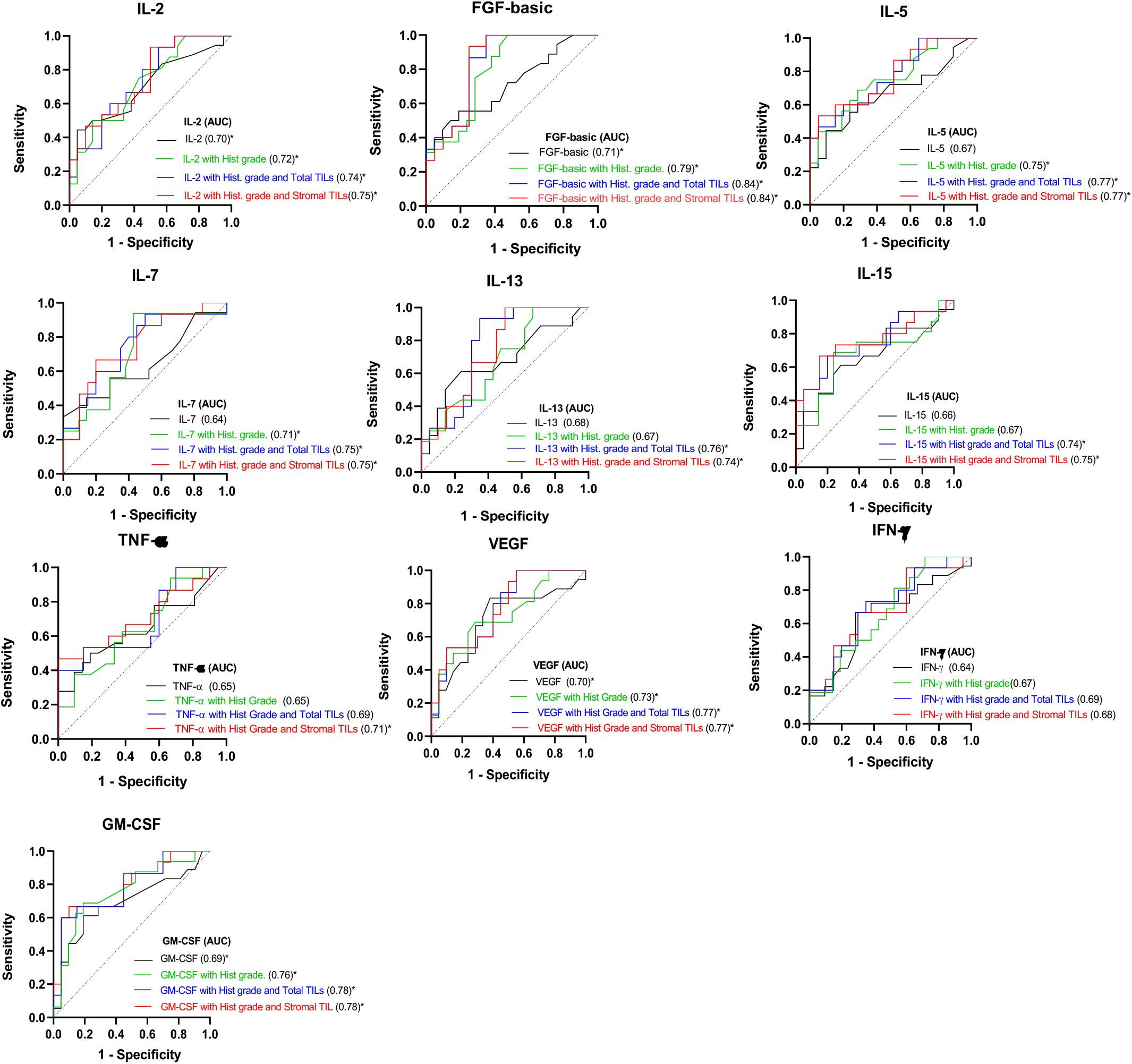
Area Under the Curve (AUC) was calculated for each cytokine (black) (univariable analysis) and in combination with histological grade (green), histological grade and Total TILs (blue) and histological grade and Stromal TILs (red) (multivariable analysis) (*p<0.05).

**Supplementary Figure 4.**
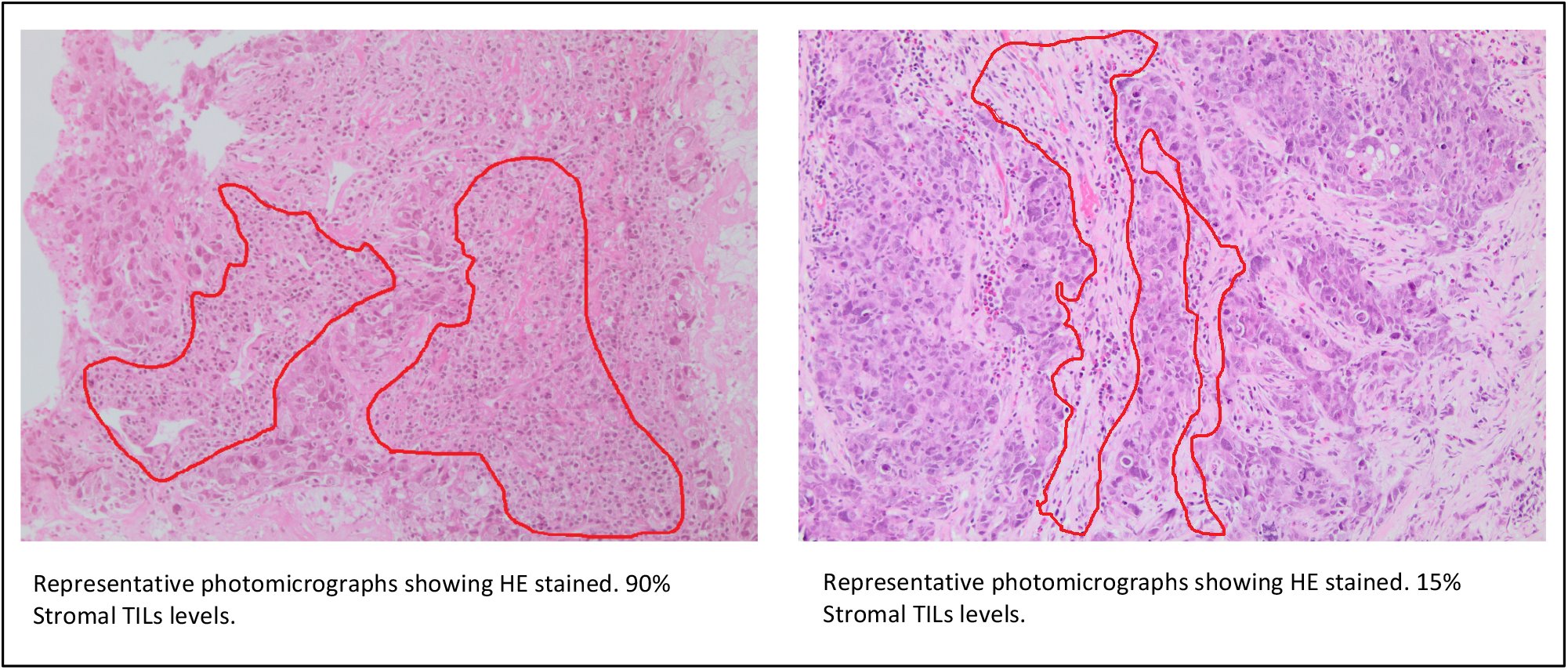
Representative Hematoxylin & Eosin stained slides from formalin fixed and paraffin embedded (FFPE) tissue blocks of core needle breast biopsies. One showing a high percentage of stromal tumor infiltrating lymphocytes (TILs) and one showing a low percentage of stromal TILs. The analysis was performed according to the recommendations by an International TILs Working Group 2014.

## Notes

### Competing Interest Statement

The authors have declared no competing interest.

### Author Declarations

This study was approved by the local Ethics Committee. A written informed consent was obtained from each patient at the time of diagnosis and before the initiation of any procedure.

